# Can MRI-based multivariate gray matter volumetric distance predict motor progression and classify slow versus fast progressors in Parkinson’s disease?

**DOI:** 10.1101/2022.07.25.22278012

**Authors:** Anupa A Vijayakumari, Hubert H Fernandez, Benjamin L Walter

## Abstract

**Introduction:** While Parkinson’s disease (PD) related neurodegeneration is associated with structural changes in the brain, magnetic resonance imaging (MRI) has not been helpful in diagnosing PD or predicting the progression of motor symptoms. In this study, we aimed to develop a structural MRI-based biomarker to predict the rate of progression of motor symptoms and to classify patients based on the symptom severity (i.e. slow vs. fast progressors) in the early stages of PD.

**Methods:** The study included 59 patients with PD (n=40 for the primary analysis, 19 for the validation analysis), and 55 healthy controls with structural MRI from the Parkinson’s Progression Markers Initiative (PPMI) database. We developed a patient-specific multivariate gray matter volumetric distance using Mahalanobis distance (M_GMV_) to investigate the changes in M_GMV_ over time using longitudinal linear mixed-effect model, its potential as a biomarker to predict the rate of progression of motor function (MDS-UPDRS-part III) using multiple linear regression model, and classification of patients based on symptom severity using machine learning (ML).

**Results:** M_GMV_ at BL significantly predicted changes in motor severity (p<0.05) and a trend level increase in M_GMV_ over time (p = 0.09) were noted. We obtained 85% accuracy in discriminating patients according to their symptom severity, and on an independent test cohort, an accuracy of 90% was achieved.

**Conclusions:** We identified a promising structural MRI-based biomarker for predicting the rate of progression of motor symptoms and classification of patients based on motor symptom severity.

## 1. Introduction

Parkinson’s disease (PD) is a chronic, progressive neurodegenerative disease characterized mainly by motor deficits related to structural changes in the brain ^1^. There is a great need to identify a reliable non-invasive biomarker to predict the rate of progression of motor symptoms and capable of diagnosing PD in early stages when symptoms are subtle and non-diagnostic.

The hallmark pathophysiology of PD is the degeneration of the dopaminergic neurons in the substantia nigra leading to functional as well as structural alterations in the basal ganglia-thalamocortical circuits ^2, 3^ and the presence of Lewy bodies in several regions of the brain ^4, 5^. Atrophy in brain regions can be gauged macroscopically using structural magnetic resonance imaging (MRI). Although routine clinical MRI is considered “normal” in PD, it has the potential to serve as a biomarker ^6^. From a clinical point of view, MRI-based biomarkers could be a compelling option since MRI is widely available, non-invasive with standardized acquisition parameters, and can be seamlessly integrated into their clinical workflow. A recently identified MRI prognostic biomarker, i.e., a PD-related atrophy score, is promising but has limited accuracy in predicting the outcomes in single patients with PD^7^. Therefore, there is an unmet demand to identify a robust imaging biomarker to monitor disease progression.

In PD, previous MRI studies have reported progressive gray matter volume (GMV) loss in cortical and subcortical structures^8-11^. In a critical review, caudate, putamen, temporal/hippocampal, frontal, and parietal areas were identified as the most consistently reported brain regions to show progressive gray matter loss in PD ^12^. As the disease advances, the motor symptoms of PD also progressively worsen over time ^13^. Prior studies investigating the possible association of GMV changes with the motor symptoms in PD have reported atrophy in various gray matter regions related to motor dysfunctions in PD ^1, 8^.

Although these studies have enhanced our understanding of the structural abnormalities and their relationship to motor symptoms in PD, firm conclusions cannot be drawn from a clinician’s point of view. Clinical relevance can be achieved only when we address the critical involvement of brain region(s) with disease progression in individual patients. This has been very challenging as the findings from previous studies carry information based on group-level analysis (i.e. comparing a group of patients with healthy controls) and not specific to individual patients. Therefore, in the clinical setting, developing a method that summarizes the heterogeneity of multiple gray matter regions into a single score in individual patients would be beneficial for clinical decision-making.

In this study, we aimed to develop a structural MRI-based biomarker based on a patient-specific summary score of GMV heterogeneity of multiple brain regions using Mahalanobis distance (MD) (referred to as multivariate gray matter volumetric distance) to predict the rate of progression of motor symptoms and to classify PD patients based on the severity of motor symptoms. To do so, initially, we aimed to examine the whole-brain GMV changes and their association with the changes in motor symptoms in PD. Later, we created a patient-specific summary score created from multiple brain regions and investigated its (1) changes over time, (2) ability to predict the rate of progression of motor symptoms, and (3) ability to classify patients based on the motor symptom severity using machine learning (ML).

## 2. Methods

### 2.1. Participants

Data used in this study was downloaded from the Parkinson’s Progression Markers Initiative (PPMI) database (www.ppmi-info.org/data) through a standard application process. For up-to-date information on the study, please visit www.ppmi-info.org. Inclusion criteria of the patients were (i) presence of asymmetric resting tremor or asymmetric bradykinesia or two of resting tremor, bradykinesia, and rigidity, (ii) evidence of positive <u>DaTscan</u> confirming the PD diagnosis, and (iii) patients untreated within at least six months of enrollment. Exclusion criteria for all participants were: (i) diagnosis of dementia or atypical PD syndromes; (ii) significant neurologic or psychiatric conditions; (iii) presence of MRI motion artifacts, field distortions, intensity inhomogeneities, or detectable brain injuries. PPMI has MRI data available at baseline, after 12, 24, and 48 months. The present study aims to examine the long-term changes in the gray matter heterogeneity and progression of motor symptoms; hence, we selected the lengthiest time period, i.e., baseline (BL) and 48 months of follow-up (48M). Motor symptoms were assessed OFF medication using part III of the Movement Disorder Society-Sponsored Revision of the Unified Parkinson’s Disease Rating Scale (MDS-UPDRS-III), ^14^ at BL and 48M. A total of 59 patients with PD and 55 age and gender-matched health controls (HC) were selected. 40 out of 59 patients with PD, who performed MRI and MDS-UPDRS-III at two time points: BL and 48M were used for primary analyses and to train the ML model. The rest of the patients (n=19) with MRI at BL and MDS-UPDRS-III at BL and 48M were used as an independent test cohort to validate the ML model. For HC, MRI data from a single time point was used. The PPMI study was approved by the Institutional Review Board of all participating sites, and written informed consent was obtained from all participants.

### 2.2. MRI data acquisition

All participants were scanned on a 3.0 T MRI scanner (Siemens, TrioTim, Germany). High-resolution T1-weighted (T1W) images were collected using magnetization-prepared rapid acquisition gradient echo sequence with the following parameters: repetition time/echo time/inversion time = 2300/3.0/900 ms, flip angle = 9°, voxel size = 1×1×1 mm, and number of slices = 176.

### 2.3. MRI data processing

Pre-processing of the MRI images was carried out using Computational Anatomy Toolbox 12 (CAT12; http://www.neuro.uni-jena.de/cat/) within SPM12 (Wellcome Department of Imaging Neuroscience Group, London, UK; http://www.fil.ion.ucl.ac.uk/spm) in MATLAB software (The Mathworks Inc.; MA, USA, R2021a). Briefly, after correcting for bias field inhomogeneity, the T1W images were segmented into gray matter (GM), white matter (WM), and cerebrospinal fluid (CSF) ^15^. Then, GM images were normalized into the standard Montreal Neurological Institute (MNI) space using Diffeomorphic Anatomic Registration Through Exponentiated Lie algebra algorithm (DARTEL) ^16^. Finally, smoothing was applied on normalized GM images with a Gaussian kernel of a full width at half maximum (FWHM) of 8 mm. For all participants, segmented images were visually inspected, and image quality ratings (IQR: combination of measurements of noise and spatial resolution) were above the “satisfactory” threshold (i.e., 75%). GMV was extracted from 86 regions of interest (ROIs) (supplementary table 1), generated from Hammers atlas (excluding corpus callosum, brain stem, and ventricular structures) ^17^ for each participant and for each time-point (for patients) using “Estimate mean values inside ROI” function in CAT12. The total intracranial volume (TIV) for each participant was extracted using the “Estimate TIV” function (TIV=GM+WM+CSF). To control for inter-subject variations in the head size, we divided GMV by TIV of each participant to obtain normalized GMV (GMV_N_).

### 2.4. Whole-brain morphometric analyses

Whole-brain voxel-based morphometric (VBM) analysis in CAT12 using spatially normalized and smoothed GM maps was examined. The differences in GMV between patients (at each time point) and HC were explored using two-tailed t-tests. A paired sample t-test was used to analyze the GMV differences between baseline and 48M in patients. Regression analysis was performed to assess whether whole-brain GMV at baseline was associated with changes in motor outcome. Age, TIV, and sex were included as covariates in the analysis with a voxel-wise threshold of p < 0.05 and an extent threshold of 50 voxels.

### 2.5. Multivariate gray matter volumetric distance

M_GMV_, multivariate gray matter volumetric distance was computed using MD ^18^ MD is a generalization of z-score in a multivariate space, which calculates patient-specific distance from the HC distribution. Age was regressed out as a covariate using the fitlm method in MATLAB, and the residuals (GMV_N_^r^) were used to compute MD as follows:

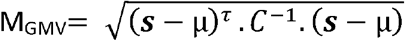

Where s represents a vector of GMV_N_^r^ observations in each ROI in a single participant, µ is the vector of average GMV_N_^r^ of each ROI calculated from HC, and C is the covariance matrix between brain regions (86 ROIs) across the HC. In general, to reliably estimate the inverse covariance matrix (*C*^-1^), at least 10 observations (HC) per GMV_N_^r^ vectors (89 ROIs) are required^19^. 50 HC were randomly selected and permuted 1000 times to compute the covariance. M_GMV_ of patients was calculated individually relative to each of the 1000 HC distributions, and the median values were reported.

### 2.6. Statistical analysis

Analyses of demographic and clinical characteristics were performed using two-tailed t-tests or Chi-square tests, as appropriate. Assumptions for normality were tested for variables using the Shapiro-Wilk test. To describe the changes in M_GMV_ over time, longitudinal linear mixed-effect models with a random participant effect and time as fixed effect were used. Changes in motor outcome were calculated by subtracting MDS-UPDRS-III scores at BL from 48M (Δ (MDS-UPDRS-III) = MDS-UPDRS-III_48M_ - MDS-UPDRS-III_BL_). Positive change scores denote a decline in motor functioning. We tested whether M_GMV_ could predict changes in motor outcome using multiple linear regression models covaried for age and sex, as follows:

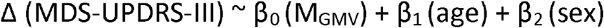

All analyses were performed using R software (version 4.1.2, https://www.r-project.org/) and p < 0.05 was considered statistically significant.

### 2.7. Machine learning for classification

We classified the patients according to Δ (UPDRS-III) into two: (i) relatively stable PD or “slow progressors,” and (ii) rapidly progressing PD or “fast progressors.” There is no clear published definition for the classification of slow vs. fast PD progressors in the literature. However, Horvath et al. defined a minimal clinically important difference (MCID) threshold of 4.63 for worsening of motor symptoms over a median interval of 6 months^20^. The present study focused on long-term outcomes, i.e., 48M, and we decided to use the median of Δ (MDS-UPDRS-III) (=7) as the threshold for the classification. Forty patients were assigned as train data, and those with Δ (MDS-UPDRS-III) < 7 were labeled as “0,” representing slow-progressors (n=19), and Δ (MDS-UPDRS-III) > 7 were labeled as “1,” representing fast-progressors (n=21). ML-based classifications were performed on PyCaret (version 2.3.6) software (https://pycaret.org/), an automated ML library in python. With M_GMV_, age, and sex as features, the logistic regression (LR) model was identified as the best model for the classification. The proposed model was evaluated using stratified 10-fold cross-validation, and patients were randomly assigned to the train and test datasets. The performance of the classifier was assessed using accuracy and area under the receiver operating characteristic curve (AUC) with specificity and sensitivity. The validity of the LR classifier model was assessed by testing it on independent test data (n=19), where each patient’s features were fed to the ML model using LR to predict the motor outcome.

## 3. Results

Clinical and demographic data were as displayed in Table 1; with no significant differences between PD and HC at baseline in age (t = 0.20; p = 0.84), or sex distribution (χ^2^ = 0.10; p = 0.74). There was a significant increase in MDS-UPDRS-III scores after 48 months (25.40 ± 11.70) compared to BL (18.50 ± 8.88) in PD (t = 2.96; p = 0.004). An increase in MDS-UPDRS-III scores was also observed in the independent test data (MDS-UPDRS-III scores after 48 months = 32.05 ± 15.40; MDS-UPDRS-III scores at BL = 21.15 ± 8.75; t = 2.66; p = 0.01). In the train data, no significant differences in the MDS-UPDRS-III scores at BL were observed between slow (20.05 ± 8.75) and fast (17.09 ± 8.96) progressors (t = 1.05; p = 0.29). After 48 months, there were significant differences in the MDS-UPDRS-III scores between slow (18.00 ± 8.64) and fast (32.04 ± 10.10) progressors (t = −4.71; p = 0.00003).

**Table 1:** Clinical and demographic data of the participants

|  | Healthy<br>controls | Patients with Parkinson's disease |  |  |  |
| --- | --- | --- | --- | --- | --- |
|  |  | Train data |  | Independent Test data |  |
|  |  | Baseline | 48 months | Baseline | 48 months |
| N | 55 | 40 | 40 | 19 | 19 |
| Age (mean ± SD) | 59.07 ± 10.93 | 58.60 ± 9.99 | 62.70 ± 9.95 | 62.52 ± 5.03 | 66.54 ± 5.03 |
| Gender (Male : Female) | 37:18 | 25:15 | 25:15 | 13:6 | 13:6 |
| Duration of illness, months (mean ± SD) | - | 5.22 ± 5.01 | 53.75 ± 5.36 | 8.78 ± 13.08 | 57.52 ± 13.94 |
| MDS-UPDRS-III total (mean ± SD) | - | 18.50 ± 8.88 | 25.40 ± 11.70 | 21.15 ± 8.75 | 32.05 ± 15.40 |

SD, standard deviation; MDS-UPDRS-III, motor section of the Movement Disorder Society-Sponsored Revision of the Unified Parkinson's Disease Rating Scale assessed in the off-medication state

### 3.1. Voxel based morphometry

Extensive gray matter volumetric reductions were noted in patients at BL and 48M compared to HC, as shown in figure 1a and supplementary table 2. GMV differences between patients at 48 months and BL were shown in figure 1b and supplementary table 3. GMV at baseline was negatively correlated with changes in MDS-UPDRS-III scores in the various brain regions, as shown in figure 1c and supplementary table 4.

**Figure 1:**
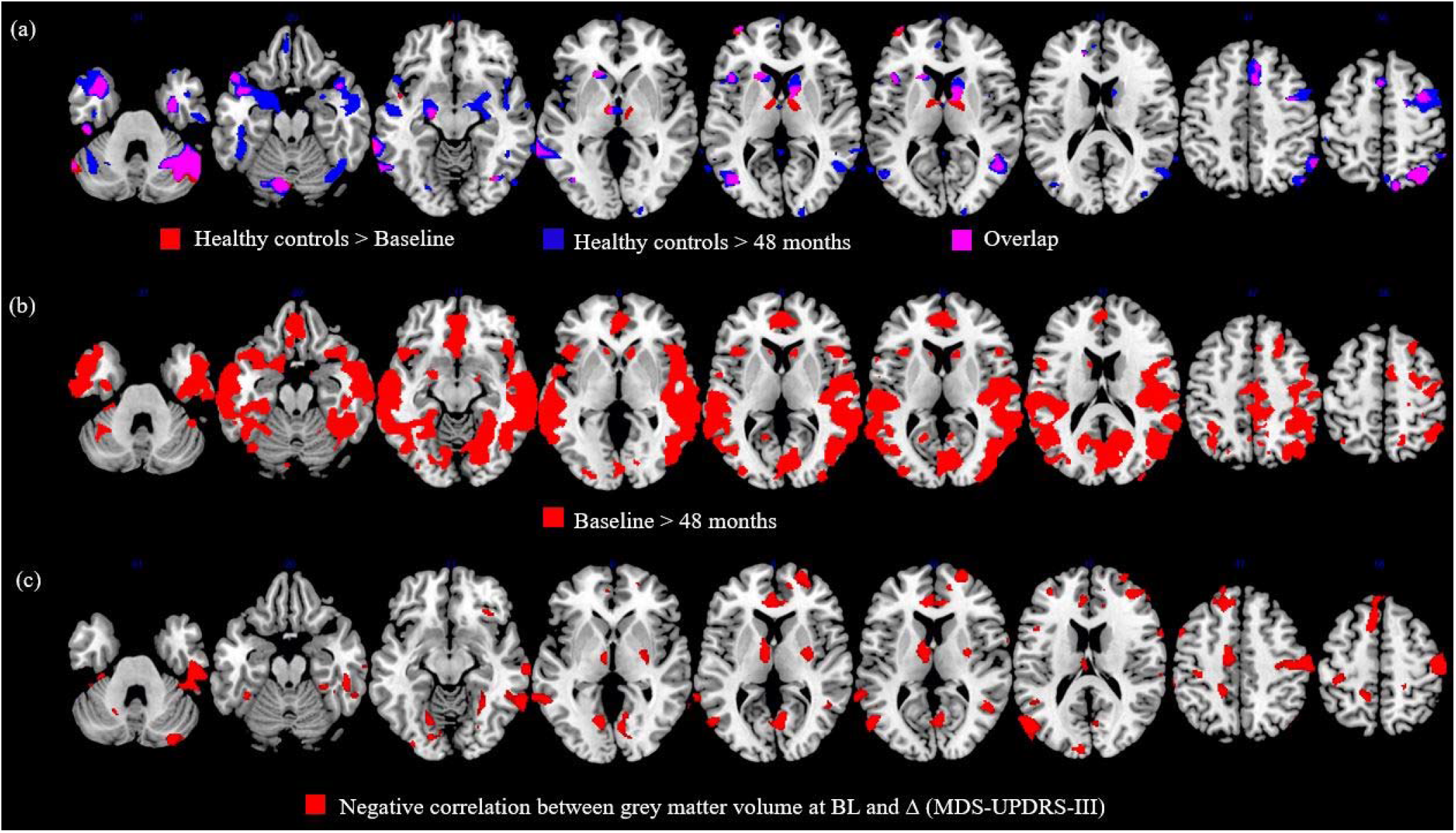
Statistical parametric maps using voxel-based morphometry analyses showing (a) differences between healthy controls and patients with PD at baseline and 48 months of follow-up, (b) differences between baseline and 48 months of follow-up, (c) negative correlation between grey matter volume at baseline and changes in MDS-UPDRS-III scores (Δ (MDS-UPDRS-III)). Results were reported using a voxel-wise threshold of p < 0.05, and an extent threshold of 50 voxels with age, total intracranial volume, and sex as covariates.

### 3.2. Longitudinal analysis of multivariate gray matter volumetric distance

Mean M_GMV_ for HC, PD at BL and PD at 48M were 8.91 ± 0.69, 9.38 ± 0.89, and 9.72 ± 0.90. Longitudinal analyses from BL to 48M exhibited an increase in M_GMV_ over time; however, it was not significant (β (95% CI) = 0.11 (−0.01 to 0.24); t = 1.67; p = 0.09) as shown in Figure 2.

**Figure 2:**
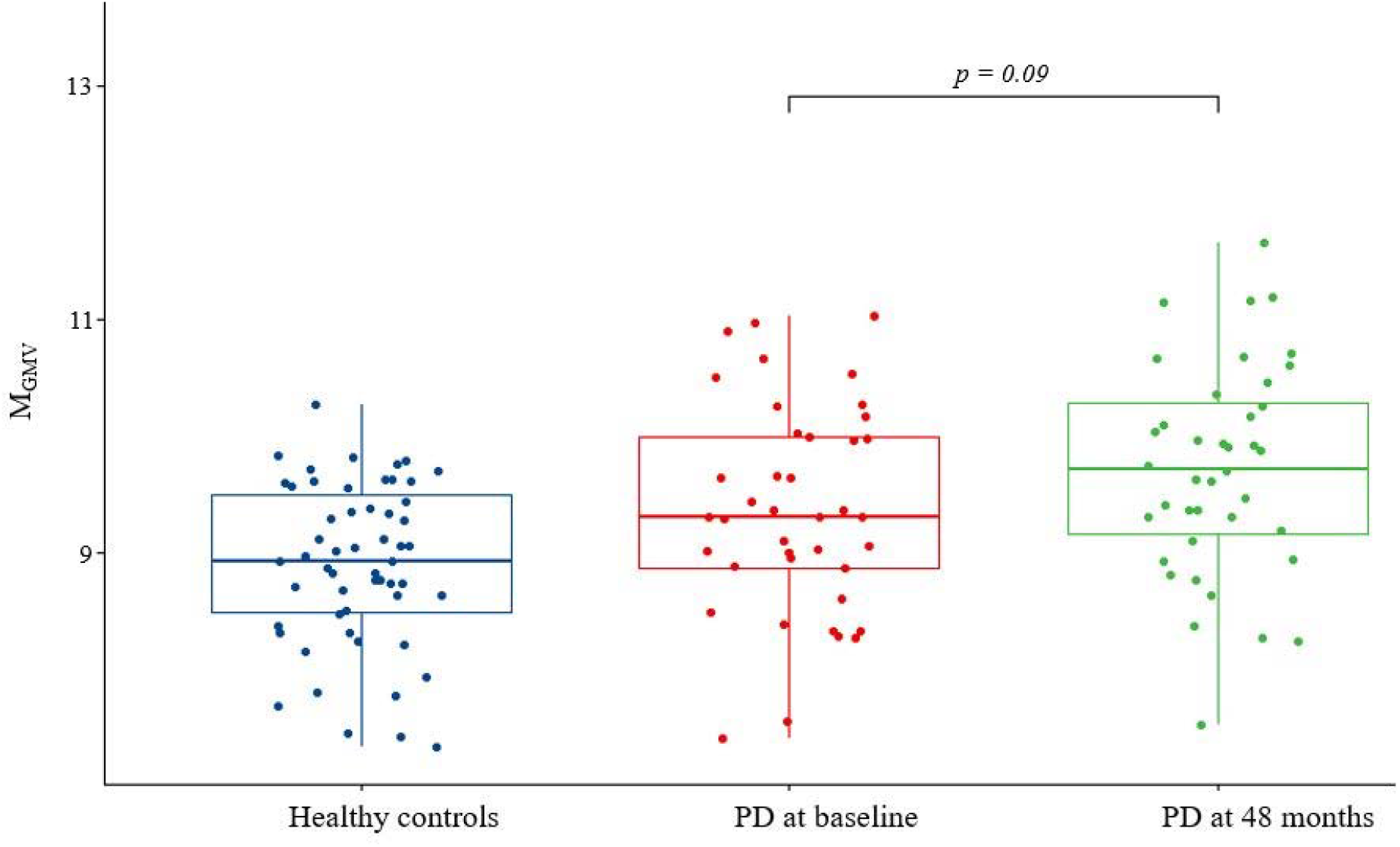
Longitudinal plot showing the changes in the patient-specific multivariate score of GM (M_GMV_) from baseline to 48-months. Healthy control data was plotted for reference.

#### 3.3. Prediction of longitudinal changes in motor severity by multivariate gray matter volumetric distance

We included baseline M_GMV_ as a predictor controlled for age and sex, with Δ (MDS-UPDRS-III) as the outcome variable in the multiple linear regression analysis. We found that M_GMV_ at baseline significantly predicted the rate of progression of motor symptoms in 40 patients with PD (β (95% CI) = 4.99 (2.18 to 7.81), adjusted R^2^= 0.28, p = 0.005). Our result implies that a one unit increase in M_GMV_ at baseline, reflects a corresponding change in the motor outcome, on an average by, 4.99 points in 48 months.

### 3.4. Classification of patients based on their motor severity using ML

Given the finding that M_GMV_ at BL could predict motor severity in 40 patients with PD, we next sought to examine if the patients could be differentiated as slow and fast progressors using ML. With these 40 patients as train data, consisting of 19 slow progressors and 21 fast progressors, LR model was tested with M_GMV_, age, at BL, and sex as features. The accuracy of the LR model for the classification of PD patients as slow and fast progressors for the train data was 85%. The AUC value reached 0.88 with a sensitivity and specificity of 80% and 91%, respectively. On independent test data comprising 8 slow and 11 fast progressors, we achieved an accuracy of 90% in classifying slow progressors from fast progressors. The AUC value was 0.90 with a specificity and sensitivity of 91%. The confusion matrices (summarizing the prediction results) for the model on the train and independent test datasets are shown in Figure 3.

**Figure 3:**
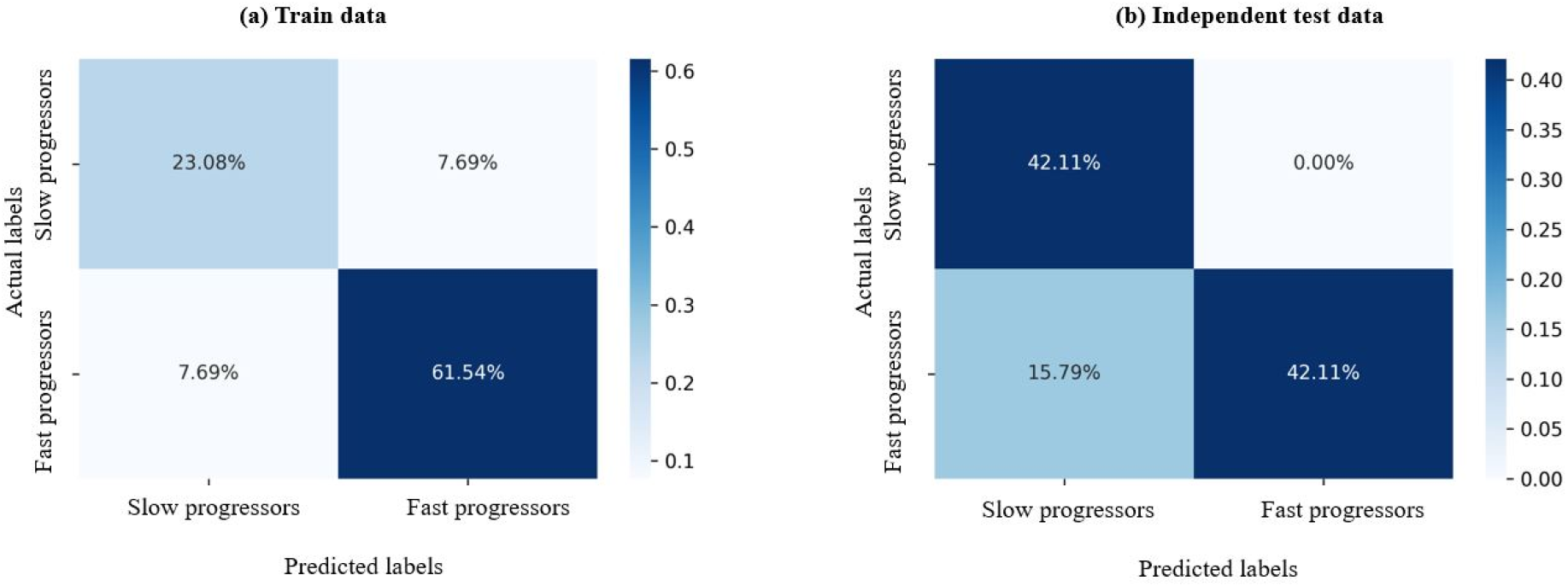
Confusion matrices depicting the performance of classifiers trained to identify slow progressors and fast progressors in (a) train data and (b) independent test data.

## 4. Discussion

In the present study, we first demonstrated grey matter volume reductions in various cortical and subcortical regions in patients with PD using whole-brain voxel-based morphometric analyses. Additionally, GMV reductions at baseline were associated with changes in motor scores. Second, we introduced a patient-specific summary score capturing the GMV heterogeneity of 86 regional gray matter volumes into a single score using MD (M_GMV_) and examined its changes over time. We observed that M_GMV_ describing the multivariate gray matter volumetric distance showed a trend towards an increase from BL to 48M. Third, M_GMV_ at BL significantly predicted the rate of progression of motor symptoms in patients with PD. Finally, we classified the patients as slow progressors and fast progressors with higher accuracy using the LR machine learning model. Thus, we developed a promising structural MRI-based biomarker for predicting the rate of progression of motor symptoms and classification of patients based on motor symptom severity.

The underlying pathology of PD involves degeneration of dopaminergic neurons in the substantia nigra, neuroinflammation, and accumulation of misfolded α-synuclein proteins as Lewy bodies and neurites in several regions of the brain ^4, 21, 22^. These processes can result in the alteration of brain morphology. Consistent with previous studies, ^10, 23, 24^ we also found gray matter volume loss in the cortical and subcortical regions in patients with PD when compared to HC and when comparing baseline and 48 months, which conform to the pathological mechanisms in PD. We also examined the association of whole-brain GMV at baseline on changes in MDS-UPDRS-III scores and observed that reduced GMV in widespread areas were associated with changes in motor scores. These findings suggest that patients diagnosed with PD have an extensive neuroanatomical pathology, which aligns with the worsening of motor symptoms.

Given the understanding that GMV changes were observed in both cortical and subcortical structures and present different cortical and subcortical atrophy patterns during PD progression^10^, we created a patient-specific summary score of GMV heterogeneity in individual patients using Mahalanobis distance, M_GMV_. MD is a promising multivariate statistical approach that combines information from numerous brain regions into a single measure, and its clinical application has been successful in traumatic brain injury, epilepsy, and autism^19, 25-28^. M_GMV_ describes the individual GMV heterogeneity by comparing one patient against a group of healthy controls. Hence, the larger the distance, the farther the patient is away from the control distribution. Longitudinal analysis of M_GMV_ demonstrated a trend level increase in the multivariate distance of patients over time, suggesting a pattern of gray matter deterioration associated with disease progression. To our best knowledge, this is the first study to apply MD of GMV in patients with PD over an extended period of time, and future studies are required to further confirm our findings.

A novel finding of this study was that M_GMV_ at baseline significantly predicted the rate of progression of motor symptoms measured using MDS-UPDRS-III. PPMI aims to identify biomarkers and define longitudinal progression biomarkers for PD cohorts using clinical and imaging features^29^. Previously proposed promising structural imaging biomarkers for disease progression and prognosis in PD were increased free water in posterior substantia nigra using diffusion MRI data ^30^ and PD-specific network atrophy pattern derived from structural MRI using Deformation-based morphometry^7^. Our results indicate that M_GMV_ could be used as a reliable imaging biomarker to support clinical decision-making in individual patients with PD. M_GMV_ can be easily obtained by processing T1-Weighted MRI in the CAT12 toolbox, which has updated atlases, including Hammers atlas and computing MD for each patient from the control population (the code for computing MD is made publicly available by Taylor et al. ^25^). Future studies adopting this framework are warranted to validate the use of M_GMV_ as a prognostic biomarker in PD.

Finally, a ML model using LR was created to discriminate between patients with slow progression and fast progression of motor symptoms. The model was trained on 40 PD patients with M_GMV_, age, and gender at baseline as features and achieved an accuracy of 85%. The discriminating ability of the model to correctly identify patients as slow and fast progressors was confirmed with an excellent AUC of 0.88. The same classifier achieved an accuracy of 90% on an independent test dataset with an AUC of 0.90. When M_GMV_ alone was used as a feature for our analysis, the accuracy of train data dropped from 85% to 60% (data not shown), indicating that demographic features such as age and sex are essential in improving the classifier’s accuracy. With the higher accuracy of our classifier on the train and test data, our models’ ability to accurately stratify patients according to their motor outcomes is very promising, and it may act as a useful prognostic classifier in clinics.

Our study has a few limitations. PD exhibits both motor and non-motor symptoms. While the present study focused on the utility of M_GMV_ in predicting and classifying patients based only on motor symptoms, future studies interrogating the capability of M_GMV_ to predict non-motor functions are warranted. We also acknowledge that our study did not measure M_GMV_ at 12 and/or 24 months (available in the PPMI database). Therefore, our understanding of the evolution of M_GMV_ and its predictability during those time periods are unknown and future work should benefit by replicating our results in a sample with data available across all time points. Although our classification results were promising, it is essential to test the performance of our classifier in a large sample size. Our results have been validated from PPMI structural MRI data acquired on a 3T Siemens TrioTrim scanner. To examine the generalizability of our proposed biomarker, future studies should be conducted on a different cohort of patients with PD using varying acquisition protocols and on different scanners.

In summary, we developed a structural MRI-based biomarker to predict the rate of progression of motor symptoms and classify patients based on motor symptom severity. Our findings are an important step towards implementing a novel imaging biomarker for the personalized treatment of Parkinson’s disease.

## Data Availability

Data used in this study was downloaded from the Parkinsons Progression Markers Initiative (PPMI) database (www.ppmi-info.org/data) through a standard application process. For up-to-date information on the study, please visit www.ppmi-info.org.

## Acknowledgments

PPMI, a public-private partnership program is funded by the Michael J. Fox Foundation for Parkinson’s Research (MJFF) with funding partners 4D Pharma, Abbvie, Acurex Therapeutics, Allergan, Amathus Therapeutics, ASAP, Avid Radiopharmaceuticals, Bial Biotech, Biogen, BioLegend, Bristol-Myers Squibb, Calico, Celgene, Dacapo Brain Science, Denali, The Edmond J. Safra Foundaiton, GE Healthcare, Genentech, GlaxoSmithKline, Golub Capital, Handl Therapeutics, Insitro, Janssen Neuroscience, Lilly, Lundbeck, Merck, Meso Scale Discovery, Neurocrine Biosciences, Pfizer, Piramal, Prevail, Roche, Sanofi Genzyme, Servier, Takeda, Teva, UCB, Verily, and Voyager Therapeutics (www.ppmi-info.org/fundingpartners).

