## SupplementaryData for "Can MRI-based multivariate gray matter volumetric distance predict motor progression and classify slow versus fast progressors in Parkinson’s disease?"

**Supplementary Table 1.** 86 brain regions of interest (ROI) from Hammers atlas. Abbreviations: L = Left, R = Right, TL = Temporal Lobe, FL = Frontal Lobe, OL = Occipital Lobe, PL = Parietal Lobe, CG = Cingulum Gyrus

| No. | ROI name |
| --- | --- |
| 1 | TL hippocampus R |
| 2 | TL hippocampus L |
| 3 | TL amygdala R |
| 4 | TL amygdala L |
| 5 | TL anterior temporal lobe medial part R |
| 6 | TL anterior temporal lobe medial part L |
| 7 | TL anterior temporal lobe lateral part R |
| 8 | TL anterior temporal lobe lateral part L |
| 9 | TL parahippocampal and ambient gyrus R |
| 10 | TL parahippocampal and ambient gyrus L |
| 11 | TL superior temporal gyrus middle part R |
| 12 | TL superior temporal gyrus middle part L |
| 13 | TL middle and inferior temporal gyrus R |
| 14 | TL middle and inferior temporal gyrus L |
| 15 | TL fusiform gyrus R |
| 16 | TL fusiform gyrus L |
| 17 | cerebellum R |
| 18 | cerebellum L |
| 19 | insula posterior long gyrus L |
| 20 | insula posterior long gyrus R |
| 21 | OL lateral remainder occipital lobe L |
| 22 | OL lateral remainder occipital lobe R |
| 23 | CG anterior cingulate gyrus L |
| 24 | CG anterior cingulate gyrus R |
| 25 | CG posterior cingulate gyrus L |
| 26 | CG posterior cingulate gyrus R |
| 27 | FL middle frontal gyrus L |
| 28 | FL middle frontal gyrus R |
| 29 | TL posterior temporal lobe L |
| 30 | TL posterior temporal lobe R |
| 31 | PL angular gyrus L |
| 32 | PL angular gyrus R |
| 33 | caudate nucleus L |
| 34 | caudate nucleus R |
| 35 | nucleus accumbens L |
| 36 | nucleus accumbens R |
| 37 | putamen L |
| 38 | putamen R |
| 39 | thalamus L |
| 40 | thalamus R |
| 41 | pallidum L |
| 42 | pallidum R |
| 43 | FL precentral gyrus L |
| 44 | FL precentral gyrus R |
| 45 | FL straight gyrus L |
| 46 | FL straight gyrus R |
| 47 | FL anterior orbital gyrus L |
| 48 | FL anterior orbital gyrus R |
| 49 | FL inferior frontal gyrus L |
| 50 | FL inferior frontal gyrus R |
| 51 | FL superior frontal gyrus L |
| 52 | FL superior frontal gyrus R |
| 53 | PL postcentral gyrus L |
| 54 | PL postcentral gyrus R |
| 55 | PL superior parietal gyrus L |
| 56 | PL superior parietal gyrus R |
| 57 | OL lingual gyrus L |
| 58 | OL lingual gyrus R |
| 59 | OL cuneus L |
| 60 | OL cuneus R |
| 61 | FL medial orbital gyrus L |
| 62 | FL medial orbital gyrus R |
| 63 | FL lateral orbital gyrus L |
| 64 | FL lateral orbital gyrus R |
| 65 | FL posterior orbital gyrus L |
| 66 | FL posterior orbital gyrus R |
| 67 | FL subgenual frontal cortex L |
| 68 | FL subgenual frontal cortex R |
| 69 | FL subcallosal area L |
| 70 | FL subcallosal area R |
| 71 | FL pre-subgenual frontal cortex L |
| 72 | FL pre-subgenual frontal cortex R |
| 73 | TL superior temporal gyrus anterior part L |
| 74 | TL superior temporal gyrus anterior part R |
| 75 | PL supramarginal gyrus L |
| 76 | PL supramarginal gyrus R |
| 77 | insula anterior short gyrus L |
| 78 | insula anterior short gyrus R |
| 79 | insula middle short gyrus L |
| 80 | insula middle short gyrus R |
| 81 | insula posterior short gyrus L |
| 82 | insula posterior short gyrus R |
| 83 | insula anterior inferior cortex L |
| 84 | insula anterior inferior cortex R |
| 85 | insula anterior long gyrus L |
| 86 | insula anterior long gyrus R |

**Supplementary Table 2.** Whole-brain grey matter regions showing differences between patients with PD and healthy controls using voxel-based morphometry analyses. Local maxima were reported, including cluster size and anatomical areas (defined in [automated anatomical labelling, AAL atlas)](https://www.sciencedirect.com/science/article/pii/S1053811919307803). Findings were reported in the Montreal Neurological Institute space (x, y, and z). Results were reported using a voxel-wise threshold of p < 0.05, and an extent threshold of 50 voxels with age, total intracranial volume, and sex as covariates. Abbreviations: L = Left, R = Right

| Anatomical area | Cluster size | MNI coordinates | |  | p value |
| --- | --- | --- | --- | --- | --- |
|  |  | **x** | **y** | **z** |  |
| *Contrast:* *Healthy controls > Patients with PD at baseline* | | | | | |
| Superior parietal gyrus_R | 888 | 34 | -66 | 60 | 0.001 |
| Superior parietal gyrus_R |  | 14 | -78 | 58 | 0.002 |
| Superior parietal gyrus_R |  | 20 | -68 | 63 | 0.011 |
| Hippocampus_L | 96 | -21 | -14 | -8 | 0.001 |
| Cerebellum_Crus2_R | 3273 | 54 | -48 | -38 | 0.001 |
| Cerebellum_Crus1_R |  | 39 | -54 | -36 | 0.002 |
| Cerebellum_7b_R |  | 42 | -39 | -45 | 0.002 |
| Caudate_L | 268 | -21 | 24 | 6 | 0.001 |
| Temporal pole: superior temporal gyrus_L | 870 | -33 | 8 | -27 | 0.001 |
| Middle temporal gyrus_L |  | -45 | -4 | -22 | 0.039 |
| Caudate_R | 561 | 9 | 3 | 6 | 0.002 |
| Thalamus_R |  | 8 | -15 | -2 | 0.035 |
| Middle frontal gyrus_L | 176 | -39 | 63 | 6 | 0.002 |
| Middle frontal gyrus_L |  | -44 | 58 | 12 | 0.004 |
| Superior frontal gyrus, medial_L | 63 | -15 | 40 | 21 | 0.003 |
| Cerebellum_Crus1_L | 151 | -57 | -58 | -28 | 0.003 |
| Temporal pole: superior temporal gyrus_L | 281 | -51 | 22 | -22 | 0.004 |
| Middle temporal gyrus_L | 643 | -63 | -45 | -4 | 0.006 |
| Middle temporal gyrus_L |  | -72 | -40 | -8 | 0.009 |
| Middle temporal gyrus_L |  | -72 | -42 | 2 | 0.012 |
| Superior frontal gyrus_R | 165 | 26 | 52 | 40 | 0.006 |
| Supplementary motor area_R | 688 | 2 | 0 | 72 | 0.006 |
| Supplementary motor area_L |  | 0 | 22 | 66 | 0.008 |
| Supplementary motor area_L |  | -3 | 16 | 60 | 0.013 |
| Superior frontal gyrus, medial orbital _L | 108 | -3 | 70 | -15 | 0.006 |
| Gyrus rectus_L |  | -9 | 54 | -16 | 0.041 |
| Middle temporal pole_R | 106 | 42 | 4 | -28 | 0.006 |
| Fusiform gyrus _R | 228 | 32 | -4 | -30 | 0.007 |
| Inferior temporal gyrus _R |  | 32 | 2 | -39 | 0.014 |
| Superior frontal gyrus, medial_R | 370 | 10 | 32 | 40 | 0.007 |
| Cingulate gyrus, mid part _R |  | 4 | 18 | 46 | 0.023 |
| Superior frontal gyrus, medial_L |  | -2 | 40 | 36 | 0.038 |
| Superior temporal gyrus_L | 207 | -50 | -4 | -6 | 0.01 |
| Precentral gyrus _R | 376 | 39 | 0 | 44 | 0.01 |
| Middle frontal gyrus_R |  | 33 | 3 | 54 | 0.012 |
| Precentral gyrus _R |  | 57 | 8 | 42 | 0.02 |
| Cerebellum_9_R | 98 | 15 | -46 | -44 | 0.01 |
| Cerebellum_Crus1_L | 432 | -10 | -72 | -22 | 0.01 |
| Cerebellum_6_L |  | -10 | -58 | -27 | 0.033 |
| Cerebellum_6_L |  | -24 | -74 | -21 | 0.041 |
| Cerebellum_8_L | 122 | -36 | -46 | -44 | 0.011 |
| Inferior temporal gyrus _L | 136 | -45 | -26 | -32 | 0.012 |
| Superior frontal gyrus_R | 259 | 20 | 3 | 68 | 0.012 |
| Superior frontal gyrus_R |  | 20 | 3 | 76 | 0.025 |
| Superior frontal gyrus_R |  | 20 | -3 | 56 | 0.027 |
| Thalamus_L | 226 | -10 | -2 | 6 | 0.012 |
| Thalamus_L |  | -9 | -9 | 0 | 0.018 |
| Thalamus_L |  | -3 | -2 | 10 | 0.028 |
| Angular gyrus _R | 237 | 54 | -72 | 36 | 0.012 |
| Angular gyrus _R |  | 46 | -63 | 38 | 0.015 |
| Precuneus_L | 175 | -6 | -48 | 68 | 0.014 |
| Middle occipital gyrus_L | 138 | -39 | -70 | 4 | 0.016 |
| Middle temporal gyrus_R | 125 | 48 | -60 | 10 | 0.02 |
| Middle temporal gyrus_R |  | 46 | -48 | 8 | 0.038 |
| Temporal pole: superior temporal gyrus_R | 82 | 44 | 12 | -16 | 0.02 |
| Inferior frontal gyrus, triangular_L | 108 | -44 | 20 | 6 | 0.025 |
| Inferior occipital lobe_R | 82 | 34 | -69 | -8 | 0.025 |
| Inferior parietal gyrus_R | 137 | 58 | -52 | 51 | 0.027 |
| Angular gyrus _R |  | 63 | -56 | 40 | 0.041 |
| *Contrast:* *Healthy controls > Patients with PD at 48 months of follow-up* | | | | | |
| Temporal pole: superior temporal gyrus_L | 6682 | -33 | 9 | -27 | <0.001 |
| Temporal pole: superior temporal gyrus_L |  | -48 | 21 | -22 | <0.001 |
| Hippocampus_L |  | -21 | -12 | -8 | <0.001 |
| Superior parietal gyrus_R | 5783 | 34 | -64 | 60 | <0.001 |
| Angular gyrus _R |  | 52 | -74 | 38 | 0.001 |
| Middle temporal gyrus_R |  | 50 | -60 | 10 | 0.001 |
| Middle temporal gyrus_R | 10454 | 44 | 4 | -27 | <0.001 |
| Cerebellum_7b_R |  | 42 | -39 | -44 | <0.001 |
| Cerebellum_Crus2_R |  | 54 | -50 | -39 | <0.001 |
| Middle temporal gyrus_L | 9426 | -63 | -45 | -4 | <0.001 |
| Middle temporal gyrus_L |  | -72 | -42 | 3 | 0.001 |
| Middle temporal gyrus_L |  | -72 | -39 | -9 | 0.001 |
| Supplementary motor area_R | 3099 | 2 | 2 | 69 | 0.001 |
| Supplementary motor area_L |  | -2 | 20 | 63 | 0.001 |
| Supplementary motor area_L |  | 0 | 10 | 72 | 0.002 |
| Hippocampus_R | 1189 | 26 | -16 | -6 | 0.001 |
| Amygdala_R |  | 27 | 3 | -15 | 0.002 |
| Hippocampus_R |  | 10 | -6 | -12 | 0.007 |
| Superior frontal gyrus, medial_L | 415 | -15 | 40 | 21 | 0.001 |
| Cingulate gyrus, anterior part_L |  | -6 | 48 | 12 | 0.01 |
| Caudate_L | 565 | -21 | 24 | 4 | 0.001 |
| Caudate_L |  | -16 | 26 | -6 | 0.006 |
| Precentral gyrus _R | 3020 | 54 | 8 | 42 | 0.002 |
| Precentral gyrus _R |  | 48 | 9 | 51 | 0.003 |
| Precentral gyrus _R |  | 40 | 0 | 44 | 0.003 |
| Caudate_R | 1740 | 9 | 4 | 8 | 0.002 |
| Thalamus_L |  | -4 | -9 | 0 | 0.015 |
| Thalamus_L |  | -2 | -4 | 9 | 0.023 |
| Inferior frontal gyrus, triangular_L | 614 | -44 | 20 | 4 | 0.004 |
| Inferior frontal gyrus, triangular_L |  | -57 | 15 | 30 | 0.012 |
| Inferior frontal gyrus, opercular _L |  | -54 | 15 | 8 | 0.021 |
| Middle frontal gyrus_L | 99 | -38 | 64 | 6 | 0.004 |
| Middle frontal gyrus_L |  | -42 | 60 | 12 | 0.023 |
| Superior frontal gyrus, medial orbital _L | 764 | -3 | 68 | -14 | 0.005 |
| Gyrus rectus_L |  | -6 | 46 | -18 | 0.007 |
| Cerebellum_9_R | 134 | 16 | -46 | -44 | 0.005 |
| Superior frontal gyrus_R | 191 | 26 | 51 | 39 | 0.006 |
| Precuneus_L | 276 | -3 | -50 | 70 | 0.007 |
| Middle occipital gyrus_L | 205 | -42 | -75 | 20 | 0.007 |
| Superior occipital lobe_R | 425 | 18 | -99 | 6 | 0.007 |
| Cuneus_R |  | 10 | -98 | 10 | 0.031 |
| Middle frontal gyrus_R | 446 | 40 | 38 | 42 | 0.008 |
| Middle frontal gyrus_R |  | 32 | 24 | 39 | 0.016 |
| Middle frontal gyrus_R |  | 38 | 52 | 32 | 0.02 |
| Inferior temporal gyrus _R | 503 | 69 | -42 | -8 | 0.008 |
| Middle temporal gyrus_R |  | 72 | -44 | 6 | 0.015 |
| Middle temporal gyrus_R |  | 63 | -45 | 6 | 0.019 |
| Vermis_4_5 | 51 | 0 | -46 | 6 | 0.012 |
| Postcentral gyrus_L | 66 | -51 | -36 | 58 | 0.013 |
| Middle occipital gyrus_R | 305 | 39 | -82 | 15 | 0.016 |
| Middle occipital gyrus_R |  | 33 | -87 | 26 | 0.026 |
| Middle occipital gyrus_R |  | 33 | -81 | 6 | 0.03 |
| Cuneus_L | 162 | -6 | -88 | 20 | 0.016 |
| Cuneus_R |  | 10 | -82 | 21 | 0.031 |
| Cerebellum_4_5_R | 66 | 26 | -24 | -30 | 0.017 |
| Rolandic operculum _R | 244 | 44 | 6 | 14 | 0.019 |
| Inferior frontal gyrus, opercular _R |  | 44 | 14 | 18 | 0.025 |
| Insula_R |  | 32 | 2 | 15 | 0.033 |
| Precentral gyrus _R | 113 | 26 | -22 | 63 | 0.02 |
| Superior frontal gyrus, medial_L | 73 | -4 | 66 | 8 | 0.021 |
| Postcentral gyrus_L | 57 | -26 | -28 | 72 | 0.021 |
| Precuneus_R | 51 | 8 | -51 | 54 | 0.027 |
| Precuneus_L | 193 | -24 | -56 | 3 | 0.03 |
| Calcarine fissure and surrounding cortex_L |  | -21 | -63 | 6 | 0.04 |
| Lingual gyrus_L |  | -16 | -52 | 3 | 0.04 |

**Supplementary Table 3.** Whole-brain grey matter regions showing differences between baseline and 48 months of follow-up using voxel-based morphometry analyses. Local maxima were reported, including cluster size and anatomical areas (defined in [automated anatomical labelling, AAL atlas)](https://www.sciencedirect.com/science/article/pii/S1053811919307803). Findings were reported in the Montreal Neurological Institute space (x, y, and z). Results were reported using a voxel-wise threshold of p < 0.05, and an extent threshold of 50 voxels with age, total intracranial volume, and sex as covariates. Abbreviations: L = Left, R = Right

| Anatomical area | Cluster size | MNI coordinates | |  | p value |
| --- | --- | --- | --- | --- | --- |
|  |  | **x** | **y** | **z** |  |
| Middle temporal gyrus_R | 86466 | 44 | -70 | 10 | <0.001 |
| Cingulate gyrus, mid part _R |  | 4 | -34 | 40 | <0.001 |
| Middle temporal gyrus_L |  | -63 | -8 | -16 | <0.001 |
| Gyrus rectus_R | 4859 | 2 | 46 | -15 | <0.001 |
| Cingulate gyrus, anterior part_R |  | 2 | 46 | 8 | <0.001 |
| Gyrus rectus_R |  | 6 | 30 | -14 | <0.001 |
| Inferior frontal gyrus, opercular _L | 329 | -56 | 9 | 16 | <0.001 |
| Inferior frontal gyrus, opercular _L |  | -46 | 9 | 26 | 0.036 |
| Superior frontal gyrus_R | 513 | 24 | 24 | 56 | <0.001 |
| Superior frontal gyrus_R |  | 22 | 18 | 46 | 0.007 |
| Inferior frontal gyrus, orbital_R | 56 | 51 | 50 | -10 | 0.001 |
| Caudate_R | 252 | 10 | 18 | 10 | 0.001 |
| Caudate_R |  | 9 | 21 | 2 | 0.003 |
| Postcentral gyrus_R | 85 | 22 | -27 | 58 | 0.003 |
| Hippocampus_R | 170 | 18 | -4 | -14 | 0.004 |
| Cerebellum_8_R | 53 | 32 | -40 | -44 | 0.005 |
| Cingulate gyrus, mid part _R | 311 | 2 | 27 | 34 | 0.006 |
| Cingulate gyrus, mid part _R |  | 6 | 16 | 42 | 0.017 |
| Supplementary motor area_R |  | 6 | 24 | 48 | 0.022 |
| Precentral gyrus _L | 117 | -44 | 0 | 40 | 0.017 |
| Precentral gyrus _L |  | -40 | 8 | 36 | 0.018 |

**Supplementary Table 4.** Whole-brain grey matter regions showing correlation between grey matter volume at baseline of patients with PD and changes in MDS-UPDRS-III scores (∆ MDS-UPDRS-III) using voxel-based morphometry analyses. Local maxima were reported, including cluster size and anatomical areas (defined in [automated anatomical labelling, AAL atlas)](https://www.sciencedirect.com/science/article/pii/S1053811919307803). Findings were reported in the Montreal Neurological Institute space (x, y, and z). Results were reported using a voxel-wise threshold of p < 0.05, and an extent threshold of 50 voxels with age, total intracranial volume, and sex as covariates. Abbreviations: L = Left, R = Right

| Anatomical area | Cluster size | MNI coordinates | |  | p value |
| --- | --- | --- | --- | --- | --- |
|  |  | **x** | **y** | **z** |  |
| Inferior temporal gyrus _R | 2481 | 54 | -27 | -24 | <0.001 |
| Middle temporal gyrus_R |  | 68 | -54 | -4 | 0.002 |
| Inferior temporal gyrus _R |  | 62 | -42 | -28 | 0.002 |
| Precuneus_L | 2715 | -10 | -54 | 75 | <0.001 |
| Postcentral gyrus_L |  | -39 | -32 | 62 | 0.007 |
| Paracentral_Lobule_L |  | -14 | -38 | 76 | 0.009 |
| Supplementary motor area_L | 1378 | -8 | 21 | 56 | <0.001 |
| Superior frontal gyrus_L |  | -14 | 36 | 45 | 0.004 |
| Superior frontal gyrus_L |  | -16 | 50 | 48 | 0.016 |
| Postcentral gyrus_R | 3262 | 52 | -20 | 63 | 0.001 |
| SupraMarginal_R |  | 64 | -18 | 46 | 0.006 |
| Postcentral gyrus_R |  | 40 | -30 | 70 | 0.007 |
| Middle frontal gyrus_R | 519 | 34 | 46 | 14 | 0.001 |
| Inferior frontal gyrus, triangular_R |  | 54 | 42 | 18 | 0.023 |
| Lingual gyrus_L | 1930 | -18 | -68 | -4 | 0.001 |
| Lingual gyrus_R |  | 6 | -72 | 3 | 0.004 |
| Lingual gyrus_R |  | 8 | -63 | 6 | 0.004 |
| Middle occipital gyrus_L | 1106 | -58 | -74 | 15 | 0.003 |
| Middle temporal gyrus_L |  | -54 | -68 | 6 | 0.011 |
| Angular gyrus _L |  | -58 | -70 | 32 | 0.011 |
| Superior frontal gyrus_R | 519 | 26 | 62 | 9 | 0.004 |
| Middle frontal gyrus_R |  | 27 | 51 | 3 | 0.006 |
| Superior frontal gyrus_R |  | 32 | 60 | 15 | 0.01 |
| Postcentral gyrus_R | 229 | 24 | -34 | 60 | 0.004 |
| Cingulate gyrus, anterior part_L | 1299 | -9 | 38 | 9 | 0.004 |
| Cingulate gyrus, anterior part_L |  | -8 | 38 | 24 | 0.005 |
| Cingulate gyrus, anterior part_R |  | 3 | 20 | 30 | 0.006 |
| Thalamus_L | 674 | -6 | -4 | 9 | 0.005 |
| Thalamus_L |  | -4 | -18 | 16 | 0.026 |
| Thalamus_L |  | -8 | -8 | -8 | 0.028 |
| Cerebellum_8_R | 158 | 28 | -56 | -44 | 0.005 |
| Cerebellum_7b_R |  | 34 | -64 | -46 | 0.034 |
| Middle frontal gyrus_L | 508 | -34 | 46 | 20 | 0.006 |
| Middle frontal gyrus_L |  | -45 | 44 | 24 | 0.018 |
| Inferior frontal gyrus, triangular_L |  | -44 | 33 | 24 | 0.025 |
| Superior frontal gyrus_R | 66 | 32 | 33 | 54 | 0.006 |
| Inferior frontal gyrus, opercular _L | 267 | -48 | 15 | 20 | 0.006 |
| Inferior frontal gyrus, triangular_L |  | -42 | 21 | 30 | 0.007 |
| Inferior parietal gyrus_L | 600 | -58 | -38 | 39 | 0.007 |
| SupraMarginal_L |  | -56 | -33 | 30 | 0.008 |
| Inferior parietal gyrus_L |  | -60 | -40 | 54 | 0.023 |
| Cingulate gyrus, mid part _L | 380 | -15 | -4 | 44 | 0.007 |
| Cingulate gyrus, mid part _L |  | -8 | -12 | 45 | 0.018 |
| Middle temporal gyrus_L | 504 | -58 | -48 | -2 | 0.008 |
| Middle temporal gyrus_L |  | -72 | -46 | -3 | 0.012 |
| Middle temporal gyrus_L |  | -70 | -46 | 8 | 0.02 |
| Cuneus_L | 176 | -9 | -92 | 16 | 0.008 |
| Fusiform gyrus _L | 353 | -38 | -45 | -16 | 0.009 |
| Cerebellum_4_5_L |  | -30 | -27 | -33 | 0.013 |
| Cerebellum_6_L |  | -38 | -36 | -27 | 0.023 |
| Inferior occipital lobe_L | 220 | -33 | -87 | -9 | 0.01 |
| Middle occipital gyrus_L |  | -46 | -90 | -8 | 0.012 |
| Precentral gyrus _L | 116 | -54 | 12 | 45 | 0.01 |
| Inferior frontal gyrus, triangular_R | 157 | 40 | 14 | 21 | 0.011 |
| Middle temporal gyrus_R | 179 | 45 | -52 | 9 | 0.011 |
| Middle temporal gyrus_R | 372 | 68 | -21 | -10 | 0.011 |
| Inferior frontal gyrus, orbital_R | 67 | 32 | 32 | -9 | 0.012 |
| Superior frontal gyrus, medial_R | 90 | 15 | 54 | 6 | 0.012 |
| Cingulate gyrus, anterior part_R |  | 12 | 45 | 26 | 0.023 |
| Putamen_R | 322 | 32 | -6 | 6 | 0.013 |
| Angular gyrus _R | 624 | 56 | -58 | 33 | 0.015 |
| Angular gyrus _R |  | 56 | -64 | 45 | 0.025 |
| Cerebellum_Crus1_R | 271 | 33 | -84 | -33 | 0.016 |
| Cerebellum_8_R | 78 | 32 | -51 | -63 | 0.018 |
| Vermis_8 | 186 | -8 | -58 | -27 | 0.019 |
| Cerebellum_6_L |  | -18 | -60 | -30 | 0.022 |
| Lingual gyrus_R | 77 | 24 | -75 | -3 | 0.021 |
| Precentral gyrus _L | 79 | -45 | -9 | 30 | 0.021 |
| Inferior frontal gyrus, opercular _L | 100 | -63 | 10 | 24 | 0.025 |
| Cerebellum_Crus2_R | 53 | 45 | -40 | -48 | 0.025 |
| Cerebellum_Crus2_R | 137 | 48 | -64 | -39 | 0.026 |
| Middle temporal gyrus_L | 141 | -48 | -51 | 14 | 0.028 |
| Inferior parietal gyrus_L | 55 | -63 | -56 | 42 | 0.029 |
| Inferior parietal gyrus_L |  | -66 | -48 | 42 | 0.031 |
| Angular gyrus _R | 55 | 48 | -74 | 44 | 0.032 |
